# Presentation and Short-term Course of New Onset Cannabis Induced Psychotic Disorder in Males

**DOI:** 10.1101/2022.01.30.22270138

**Authors:** Deepak Cyril D’Souza, Justin Raj, Suhas Ganesh, Nishant Goyal, Vidya KL, Sai Krishna Tikka, Umesh Shreekantiah, Pratima Murthy, Daya Ram, Priyamvada Sharma, Vinod K Sinha, Jose Cortes-Briones

## Abstract

**Introduction:** Cannabis use has been associated with several psychosis outcomes including acute and persistent psychosis termed Cannabis Induced Psychotic Disorder (CIPD). The clinical and cognitive profile, course of CIPD, and the extent to which it is different from psychosis unrelated to cannabis exposure (PsyNoCan) is not clear.

**Methods:** The acute presentation and short-term (∼4 weeks) course of hospitalized male patients with new onset CIPD were compared prospectively to PsyNoCan using measures of psychosis, depression, mania, memory and other cognitive processes at admission, and after 4 weeks of inpatient hospitalization. A subsample of CIPD patients were followed up after 4-6 months of discharge. Cognitive test performance was benchmarked for comparison in healthy controls and individuals with Cannabis Use Disorder.

**Results:** Compared to PsyNoCan (n=53), CIPD (n=66) had a significantly lower severity of psychotic symptoms at admission but no differences in mood symptoms. After 4 weeks of hospitalization, the CIPD group had less psychosis. While both groups had significant cognitive deficits at baseline compared to healthy controls, cognitive test performance improved to a greater extent in CIPD. Amongst 16 CIPD cases with longitudinal follow-up data, 10 relapsed with psychosis within 6 months after resuming cannabis use.

**Conclusion:** CIPD in males has a distinct presentation and short-term course, characterized by less severe psychosis, and greater resolution of psychopathology and cognitive deficits relative to PsyNoCan. Relapse of cannabis use may predict poorer long-term outcomes with greater psychotic relapses. The longer-term course, prognosis and biology of CIPD, and its presentation in females needs further study.

## Introduction

Schizophrenia (SCZ) or, as Bleuler appropriately coined, the “group of schizophrenias”, is heterogenous on a number of levels^1^ including its phenomenology and clinical manifestation, environmental risks, molecular genetics, transcriptomics, proteomics, and brain structure, function and connectivity. Several attempts have been made to identify and tease apart SCZ subtypes based on clinical manifestations, genetics, and biomarkers^2^. Converging lines of evidence suggest that exposure to cannabis increases the risk of several psychosis outcomes^reviewed in 3^. Cannabis, its principal constituent cannabinoid delta-9-tetrahydrocannabinol (THC) and synthetic CB1R receptor agonists (e.g., nabilone) have been shown to induce transient psychosis-like states in controlled laboratory studies^4^. Beyond these self-limited psychosis-like states, are persistent psychosis syndromes that occur with exposure to cannabis and synthetic cannabinoids, but which outlast the period of intoxication and warrant clinical intervention^reviewed by5^.

Both, the International Classification of Disease (ICD)-11 and the Diagnostic and Statistical Manual (DSM-5) recognize cannabis induced psychotic disorder (CIPD) as a diagnostic entity (6C41.6 or 292.9) with the caveat that in order to diagnose it, SCZ must be ruled out. Distinguishing CIPD from SCZ remains challenging given the heterogeneity of SCZ, as discussed above. Previous attempts to characterize CIPD that relied on clinical features failed to identify any signature characteristics^6, 7^. Chopra described the clinical presentation of a series of hospitalized cases of cannabis related psychosis in India^8^. The psychosis was typically preceded by ingestion of large doses of cannabis and was characterized by hallucinations, paranoia, delusions, depersonalization, emotional lability, amnesia, confusion, and disorientation. Others have described a predominantly affective psychosis characterized by unusual thought content, excitement, grandiosity, hallucinations, uncooperativeness, and cognitive dysfunction which improves with abstinence from cannabis.

The course of psychosis related to cannabis is intriguing. While according to the ICD-10 and DSM-5, CIPD resolves with abstinence from cannabis, several studies report that up to 50% of CIPD cases develop chronic mental disorders such as schizophrenia or related disorders. Furthermore, this risk of developing SCZ remained elevated for 10-15 years after the initial diagnosis of CIPD, suggesting that a large proportion develop or convert to a chronic psychotic disorder rather than initial misdiagnoses^9^. This raises the intriguing possibility of a cannabis related chronic psychotic syndrome that is currently lumped under “schizophrenia”.

The existing literature^6-8^ characterizing CIPD is sparse and consists mostly of case reports/series of CIPD which when viewed collectively, has several limitations. In many studies, preexisting psychosis disorder unrelated to cannabis exposure was not conclusively excluded. Most studies relied on self-reported cannabis exposure without drug testing. Likewise, the amount of cannabis use preceding the psychotic episode was not quantified, limiting assessment of dose-response. Since most studies included individuals who co-use other drugs, it is unclear to what extent, the other drug (e.g., stimulants) contributed to the development of psychosis. Most studies focused on positive symptoms, did not employ standardized measures, and did not systematically assess negative symptoms or cognitive function. Lastly, few studies have prospectively characterized the phenomenology of CIPD.

Recent changes in the cannabis landscape have raised concerns about possible increases in psychosis outcomes. The legalization of “medical” and recreational cannabis use continues to spread globally^10^. The potency (THC content) of cannabis has increased^11^. In addition to cannabis flower and hashish, a growing array of cannabis-based products and formulations are now commercially available including high-potency cannabis extracts, edibles, and beverages. With commercialization, increased access to cheaper and more potent cannabis will increase use among current users^12^. Hjorthøj et al., reported a doubling in the incidence of CIPD between 2006 and 2016 in Denmark coincident with an increase in the potency of cannabis^13^. Hjorthøj et al., noted that rates of CIPD increased independent of the increase in dually diagnosed patients i.e., comorbid cannabis use by patients with established schizophrenia.

The primary goal of this study was to characterize cases of cannabis positive psychosis vs. cannabis negative psychosis hospitalized for the first time, at admission and after 4 weeks of hospitalization using cognitive, behavioral, and toxicological measures. We further aimed to explore the relationship between recurrence of cannabis use and a relapse of psychosis in the early post-discharge follow-up period.

## Methods

### General Study Design (Figure S1)

Prospective, longitudinal observational study to characterize psychosis related to cannabis.

### Subjects (Table S1)

The primary groups of interest were individuals hospitalized for psychosis with or without exposure to cannabis. Cases of CIPD were defined as those hospitalized for the first time for psychosis positive for cannabis determined by self-report, collateral report and urine toxicology (positive for THC-COOH). Psychosis controls (PsyNoCan) were defined as those hospitalized for psychosis but without exposure to cannabis.

In order to benchmark the cognitive and EEG outcomes (reported elsewhere), cannabis controls who were hospitalized at CIP for ICD-10 Cannabis Use Disorder but without psychosis and healthy controls without psychiatric or substance use disorders from the community, were recruited.

### Regulatory

This study was approved by the Ethics Committee of the Central Institute of Psychiatry (C.I.P), Ranchi, India and the Ethics Committee of the National Institute of Mental Health and Neurosciences (NIMHANS), Bengaluru, India, the Human Investigation Committee of Yale University School of Medicine, New Haven, USA, and the Indian Council of Medical Research (ICMR).

### Recruitment

Given the challenges to studying large numbers of cases of psychosis related to cannabis the study was conducted at the Central Institute of Psychiatry (CIP), Ranchi, India a tertiary care Central Governmental supported psychiatric facility that serves a catchment area of ∼200 million people. In 2018, CIP logged >90000 outpatient visits, and ∼4000 inpatient admissions. Their outpatient department serves ∼100 new patients and ∼350 follow ups every day. The hospital sees a steady stream of cases of psychosis related to cannabis that occur throughout the year and also cases that occur in the context of a number of annually recurring festivals (e.g., Holi), during which cannabis is widely consumed. We capitalized on the large numbers of cases of psychosis with and without exposure to cannabis to recruit for the study.

### Sampling strategy (Figure S2)

Purposive sampling was used to recruit CIPD cases and convenience sampling was used to recruit matched PsyNoCan controls. An independent clinical team of psychiatrists established diagnoses were established and then referred potential participants to the research team to initiate recruitment and screening as described in CONSORT chart (Figure S2).

### Inclusion/Exclusion Criteria (Table S2 & S3)

ICD-10 Cannabis induced psychosis (CIPD) with a history of cannabis exposure confirmed by urine toxicology or psychosis with a history of cannabis exposure (PsyNoCan) confirmed by urine toxicology. Common criteria: 1) men, 2) 18-60 years, 3) requiring hospitalization, 4) able to provide informed consent in English or Hindi, 5) absence of unstable medical or neurological conditions (including seizure disorder) that may interfere with the safety of the study or may confound the interpretation of study results, 7) absence of substance use other than tobacco, and cannabis in cases 6) urine toxicology negative for illegal drugs (other than cannabis in cases).

### Schedule of Procedures (Table S4 & S5)

Once cases, and psychosis controls were hospitalized, the research team obtained written informed consent and initiated the screening process. To accommodate potential participants who were too uncooperative or disorganized on the day of hospitalization, research procedures allowance was given to initiate procedure within ∼3 days from the day of hospitalization. Importantly the first set of assessments were not conducted while subjects were under the acute influence of cannabis. The clinical chart was reviewed and the participants’ family members who accompanied the participant were also interviewed to obtain collateral information.

### Outcome Measures (Table S6)

Cannabis exposure was established with combination of self-report, collateral report from the family, a structured interview, immediate (real-time) urine toxicology and later quantification of THC-COOH in stored urine samples. A calendar-based approach, the modified time line follow-back (TLFB), was used to estimate last month drug use, including cannabis^14^. Diagnosis was established using the Diagnostic Interview for Genetic Studies (DIGS), a structured interview for psychiatric disorders which has been validated for psychiatric research in India^15^.

Psychosis was assessed using the Positive and Negative Syndrome Scale (PANSS)^16^. Depression was assessed using the Calgary Depression Rating Scale^**17**^. Mania was assessed using the Young Mania Rating Scale^18^.

The CogState Schizophrenia Battery was used to assess speed of processing, attention/vigilance, working memory, visual learning, verbal learning, reasoning/problem solving, and social cognition, domains that are recommended by the MATRICS initiative^19, 20^. It is rapid (35 minutes), it uses culture-neutral stimuli (playing cards) and is computerized. All ratings and cognitive tests were administered by a trained Masters’ level rater. Verbal memory and recall were assessed using the modified Hopkins Verbal Learning Test (HVLT)^15^.

### Urine Toxicology

Urine was tested for 10 drugs including THC-COOH in real time using immunoassay PreScreen Plus Dip Card Urine. A random set of samples from each group were subjected to quantification of urinary carboxy-THC using Liquid-Chromatography Mass-Spectrometry (LC-MS) at the toxicology lab at NIMHANS.

## Statistical Analysis

The distributions of demographic, clinical and cognitive variables were examined visually for deviations from normality using histogram, box and whisker plot and tested using Kolmogorov-Smirnov statistics. Baseline comparisons between CIPD and PsyNoCan were performed using Student’s t-test. Longitudinal change in clinical and cognitive outcomes over 4 weeks of hospitalization were examined using linear mixed models to examine the effect group, time and group*time interaction as fixed factors and subject ID as a random factor. Influence of differences in demographic variables on clinical and cognitive outcomes were explored within the model by including these variables as covariates. Mixed models offer the advantage of greater flexibility in fitting correlated factor structures in repeated measures and are optimal when data are missing at random. Bonferroni corrections were applied for multiple comparisons within outcome domains. The relationship between change in clinical variables and change in cognitive variables was visually examined with scatter plots and was measured using Pearson’s correlation coefficient. Data were analyzed in R version 4.0.1 (R: A language and environment for statistical computing) using packages Tidyverse, lme4, and lmerTest.

## Results

### Sample Demographics (Table 1)

A total of 66 patients with CIPD and 53 with PsyNoCan were recruited. All participants in this study were men. Relative to the PsyNoCan, the CIPD group was younger, had fewer years of education, had a shorter duration of illness and were less likely to have a history of psychiatric illness.

**Table 1:** Demographics.

| Variable | CIPD | PsyNoCan | Statistics |
| --- | --- | --- | --- |
| Age in years: mean (SD) | 26.82<br>(6.95) | 29.96<br>(6.14) | t = -2.6,<br>p = .01 |
| Sex: male % | 100 | 100 | NS |
| Years of education: mean (SD) | 8.89<br>(3.83) | 10.75<br>(3.19) | t = -2.9,<br>p = 0.005 |
| Duration of illness in month: mean (SD) | 2.08<br>(1.96) | 47.18<br>(40.77) | t = -7.7,<br>p < 0.0001 |
| Significant past psych history: n (%) | 8<br>(12.1) | 23<br>(43.4) | Chi = 13.34,<br>p = 0.0003 |
| Lifetime cannabis use: n (%) | 66 (100) | 0 (0) |  |
| Age of first use: mean (SD) | 19.02 (6.27) | NA |  |
| Duration of use in years: median (IQR) | 8 (7) | NA |  |
| Most common pattern of use | Smoking | NA |  |
| Time since last use in hours: median (IQR) | 24<br>(24) | NA |  |
| Total lifetime use: ~# of joints: median (IQR) | 730<br>(2920) | NA |  |
| Urinary THC-COOH concentration ng/dl: range<br>(median) | 44.16 – 3762.68<br>(64.79) | NA |  |

#### Cannabis use profile

Every participant in the CIPD group had self-reported history of cannabis use, a positive urine toxicology for cannabis and last cannabis use 24 hours prior to hospitalization. There was 100% concordance between screening and quantification assays. The mean (SD) age of onset of cannabis use in this group was 19.32 (6.53) years and had used approximately a median of 730 joints of cannabis. Smoking was the most common mode of use either joints or *chillums* (a traditional pipe for smoking cannabis: Figure S3) with 65% of the sample having reported mixing tobacco along with cannabis. Cannabis was also consumed in the form of edibles (Figure S4). There was no self-reported history of cannabis use in the PsyNoCan group which was corroborated by family members and by negative urine toxicology. There was no self-reported synthetic cannabinoid use in both groups, and urine toxicology for any other drugs was negative in both groups. Being a drug-free hospital campus, during the hospitalization, participants were not exposed to cannabis as confirmed by urine toxicology.

### Symptom profile of CIPD in comparison with PsyNoCan

#### Admission (Figure 1)

Relative to PsyNoCan, the CIPD group had significantly lower PANSS total, positive symptoms, negative symptoms, general symptoms (all ps < 0.0001). There were no significant differences in the severity of depression or mania at the time of admission.

**Figure 1.**
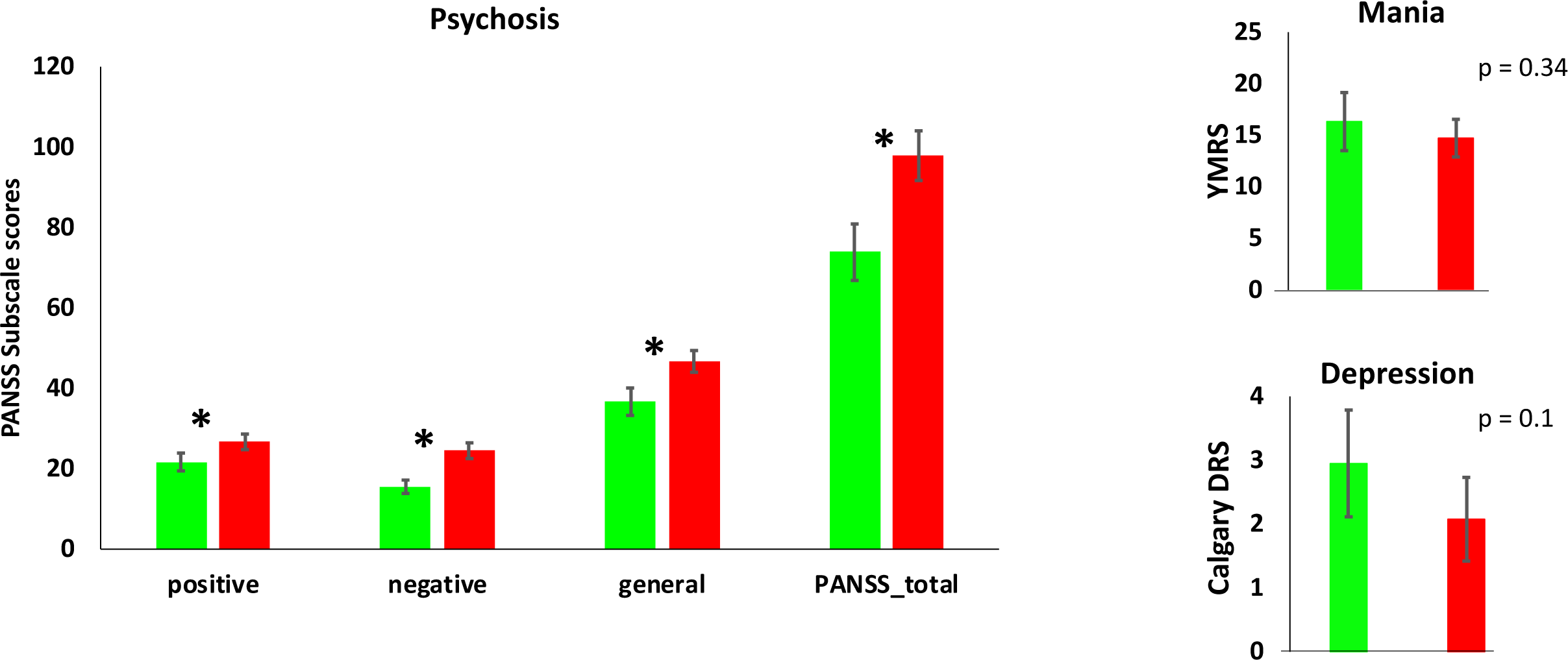
Psychosis, Depression and Mania at Admission. 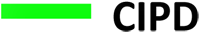 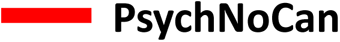 Differences in PANSS (Positive and Negative Syndrome Scale), YMRS (Young Mania Rating Scale) and CDRS (Calgary Depression Rating Scale) scores at Admission to the hospital between CIPD and PsychNoCan. Means and +/- 2SE * p <0.0001

#### Over 4-week hospitalization (Figure 2, Table 2)

At 4 weeks, compared to the PsyNoCan group, CIPD had significantly lower total PANSS scores and on the positive, negative, general PANSS subscales (all ps < 0.0001). Both CIPD and PsyNoCan groups had significant improvement in each of the PANSS domain symptom scores over four weeks of treatment but there were no significant group*time interactions on any of the measures.

**Table 2:** Group Differences in Psychosis, Mania, & Depression Over 4 weeks.

|  |  | CIPD |  | PsychNoCan |  | Statistics for change over 4 weeks |
| --- | --- | --- | --- | --- | --- | --- |
|  |  | Admission | 4 weeks | Admission | 4 weeks |  |
| PANSS | Total score | 73.88<br>(27.21) | 47.5<br>(17.37) | 97.88<br>(22.21) | 74.95<br>(23.85) | Group: 23.65 (<0.0001)<br>Time: -27.74 (<0.0001)<br>Group x time: -2.88 (0.62) |
|  | Positive | 21.68<br>(8.62) | 11.59<br>(5.04) | 26.67<br>(6.99) | 17.47<br>(6.82) | Group: 4.93 (0.0001)<br>Time: -10.45 (<0.0001)<br>Group x time: -0.55 (0.77) |
|  | Negative | 15.52<br>(6.46) | 10.98<br>(4.00) | 24.5<br>(7.08) | 20.39<br>(7.31) | Group: 8.87 (<0.0001)<br>Time: -4.87 (<0.0001)<br>Group x time: -1.27 (0.42) |
|  | General | 36.68<br>(13.16) | 24.93<br>(8.81) | 46.71<br>(9.71) | 37.08<br>(10.95) | Group: 9.85 (<0.0001)<br>Time: -12.45 (<0.0001)<br>Group x time: -1.01 (0.72) |
| YMRS |  | 16.33<br>(10.92) | 2.77<br>(3.21) | 14.73<br>(6.58) | 4.17<br>(5.35) | Group: -1.61 (0.25)<br>Time: -13.56 (<0.0001)<br>Group x time: 2.98 (0.15) |
| CDRS |  | 2.95<br>(3.24) | 0.46<br>(0.99) | 2.08<br>(2.37) | 0.33<br>(0.93) | Group: -0.88 (0.04)<br>Time: -2.5 (<0.0001)<br>Group x time: 0.73 (0.19) |
| PANSS: Positive and Negative Syndrome Scale; YMRS: Young Mania Rating Scale; CDRS: Calgary Depression Rating Scale |  |  |  |  |  |  |

**Figure 2.**
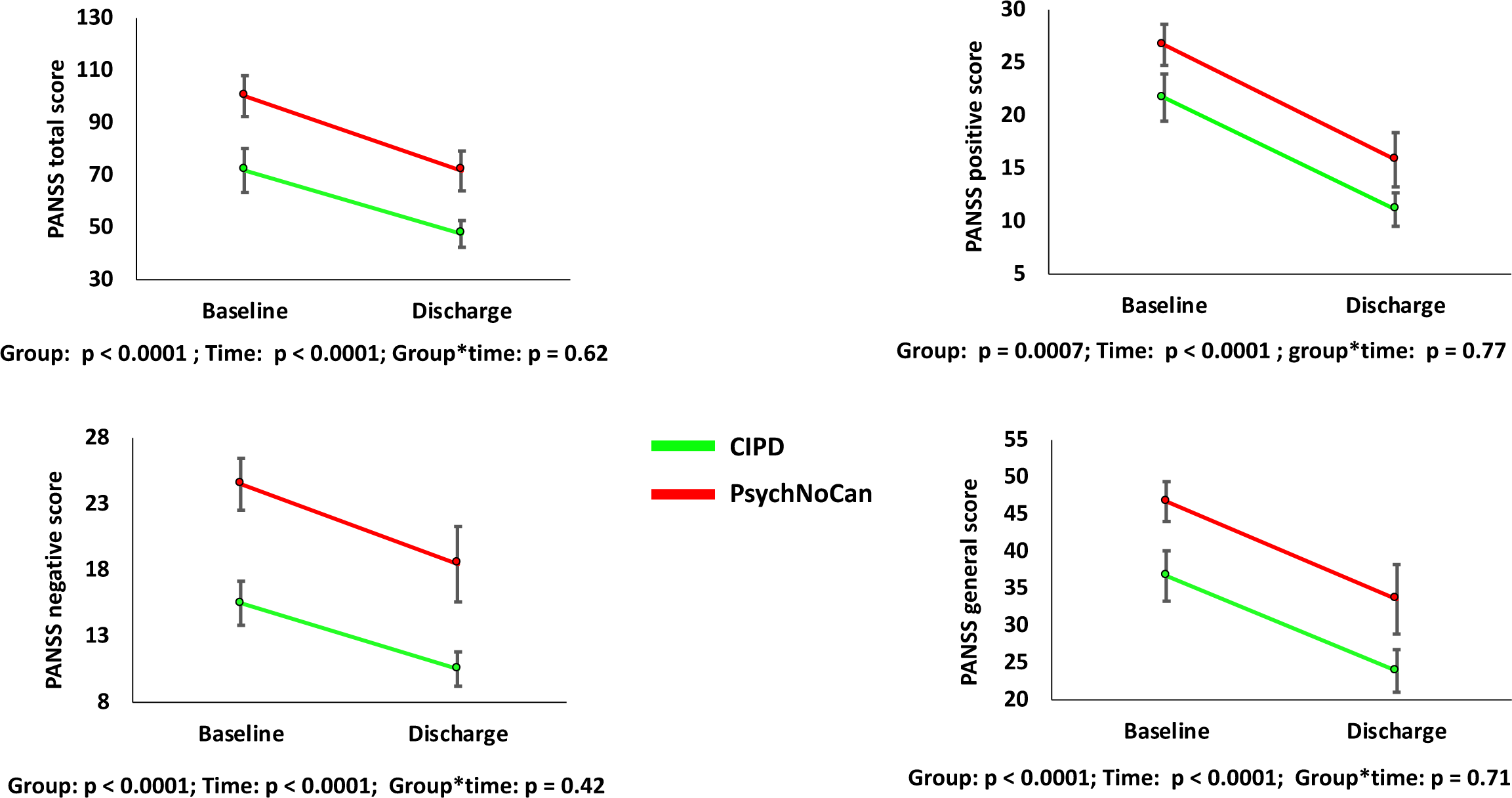
Change in Psychosis Symptoms Over 4 Weeks. 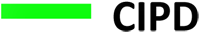 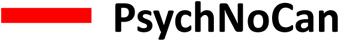 Change in PANSS (Positive and Negative Syndrome Scale) Total, Positive, Negative and General Subscale Scores from admission to the hospital to ∼ 4 weeks of inpatient hospitalization. Means and +/- 2SE

### Cognitive Performance of CIPD in comparison with PsyNoCan

#### Admission (Table 3)

There were no statistically significant differences between CIPD and PsyNoCan groups in performance on any of the cognitive tasks at admission.

**Table 3:** Group Differences in Cognitive Test Performance.

|  |  | CIPD |  | PsyNoCan |  | Statistics<br>(p value) |
| --- | --- | --- | --- | --- | --- | --- |
|  |  | Admission | 4 weeks | Admission | 4 weeks |  |
| Hopkins Verbal Learning<br>Test (HVLТ) | Immediate Recall | -0.51 | -0.16 | -0.18 | -0.36 | Group: 1.26 (0.2), Time: 1.71 (0.002),<br>Group x time: -2.12 (0.008) |
|  | Delayed Recall | -0.49 | -0.07 | -0.28 | -0.12 | Group: 0.27 (0.46), Time: 0.8<br>(0.0006) Group x time: -0.29 (0.38) |
|  | Cued Recall | -0.72 | -0.23 | 0.004 | -0.05 | Group: 1.43 (0.0003), Time: 1.04<br>(<0.0001),<br>Group x time: -1.07 (0.004) |
|  | Recognition Recall | -0.42 | -0.14 | -0.12 | -0.5 | Group: 0.53 (0.16), Time: 0.55 (0.03),<br>Group x time: -1.23 (0.001) |
| Cogstate Schizophrenia Battery | Composite score | -0.26 | 0.04 | -0.04 | -0.06 | Group: 0.19 (0.13), Time:0.33<br>(0.002),<br>Group x time: -0.34 (0.025) |
|  | Identification | -0.16 | 0.09 | -0.17 | -0.06 | Group: 0.004 (0.84), Time: -0.02<br>(0.16),<br>Group x time: 0.01 (0.63) |
|  | Detection | -0.32 | -0.11 | 0.03 | 0.14 | Group: -0.04 (0.08), Time: -0.03<br>(0.21),<br>Group x time: -0.1 (0.76) |
|  | One back test | -0.27 | 0.22 | -0.05 | 0.06 | Group: -0.03 (0.32), Time: -0.06<br>(0.01),<br>Group x time: -0.05 (0.19) |
|  | One card learning | -0.29 | 0.03 | -0.16 | 0.04 | Group: 0.01 (0.72), Time: 0.06 (0.02),<br>Group x time: -0.01 (0.67) |
|  | **Groton maze learning\*** | -0.31 | 0.12 | 0.06 | -0.31 | Group: -31.65 (0.09), Time: -41.53 (0.008),<br>Group x time: 69.42 (0.003) |
|  | **Groton maze recall** | -0.16 | 0.2 | -0.2 | -0.39 | Group: 0.92 (0.78), Time: -5.57 (0.05),<br>Group x time: 8.18 (0.049) |
|  | **Two-back test** | -0.45 | -0.08 | -0.05 | -0.07 | Group: 0.08 (0.08), Time: 0.1 (0.007),<br>Group x time: -0.09 *0.08) |
|  | **Continuous paired associate learning** | -0.26 | -0.31 | 0.1 | -0.06 | Group: -26.93 (0.13), Time: -3.4 (0.81),<br>Group x time: 9.47 (0.64) |
|  | **Social cognition** | -0.31 | -0.11 | 0.04 | -0.27 | Group: 0.09 (0.08), Time: 0.06 (0.12),<br>Group x time: -0.13 (0.02) |

Values transformed and centered to mean in the entire sample such that higher scores represent better performance on each task. Raw scores were used in the statistical analyses. Cogstate Battery: <after correction for multiple comparisons, individual tasks p values < 0.005 (0.05/9) were considered significant.

#### After 4-weeks hospitalization (Figure 3; Table 3)

After ∼4 weeks of inpatient treatment, the cognitive performance on multiple tasks significantly improved in the CIPD compared to PsyNoCan group. Statistically significant group*time interactions were noted on the domains of verbal learning (beta = -2.12, SE = 0.78, p = 0.008), the composite score of the Cogstate Schizophrenia Battery group x time: (beta = -.34, SE = 0.15, p = 0.025), visuo-spatial learning (group x time: beta = 69.42, SE = 22.58 (0.003) indicative of greater improvement in CIPD compared to PsyNoCan over 4 weeks. Furthermore, trend level differences of greater improvement in CIPD on visuo-spatial recall (beta = 8.18, SE = 4.1, p = 0.049) and social emotional cognition (beta = -0.14, SE = 0.06, p = 0.02) were noted (Table S5).

**Figure 3.**
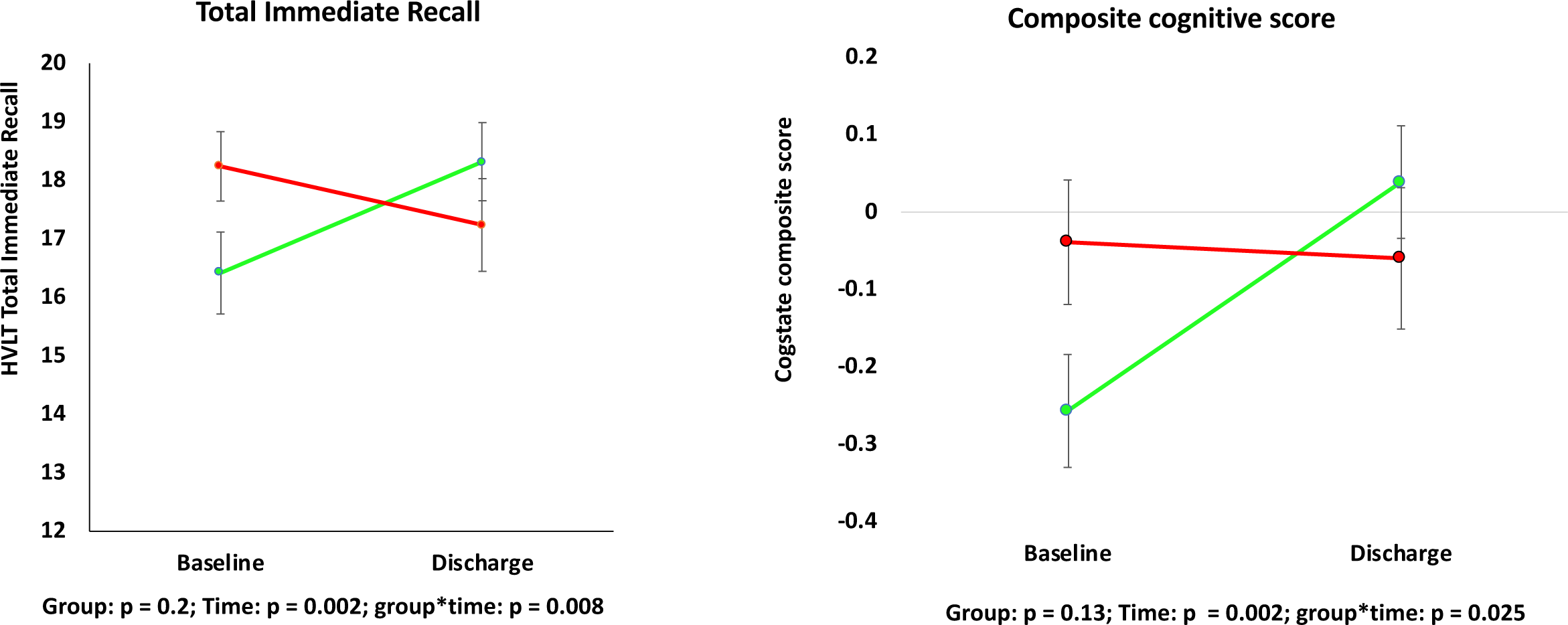
Group differences in Cognition – Admission to 4 weeks. 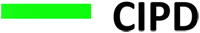 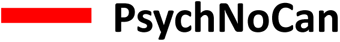 Change in Total Immediate Recall HVLT (Hopkins Verbal Learning Test), and Composite Score of the Cogstate Schizophrenia Battery from admission to the hospital to ∼ 4 weeks of inpatient hospitalization. Means and +/- 2SE

### Relationship between changes in symptoms and cognition

There was no correlation between improvement in cognitive symptoms measured as change in Cogstate composite score and improvement in psychosis symptoms measured as change in PANSS total score (r = -0.04, p = 0.83).

### Short-term Longitudinal Course in CIPD (Figure 4)

Follow-up data was collected ∼4-6 months after the index hospitalization in a subsample of CIPD participants (n=16). Within 6 months of the index hospitalization, 10 CIPD participants were re-hospitalized for a relapse of psychosis related to the resumption of cannabis use. The latter was confirmed by urine toxicology, self-report, and collateral information provided by the family. Among the 6 participants who did not relapse, only one tested positive urinary THC-COOH. In this preliminary and exploratory analysis, recurrence of cannabis use was significantly associated with recurrence of psychosis (Fisher test, p = 0.001). Additionally, of the 10 CIPD cases who had a psychosis relapse 3 patients were free from psychotic symptoms for 4 months after discharge until they resumed cannabis use.

**Figure 4:**
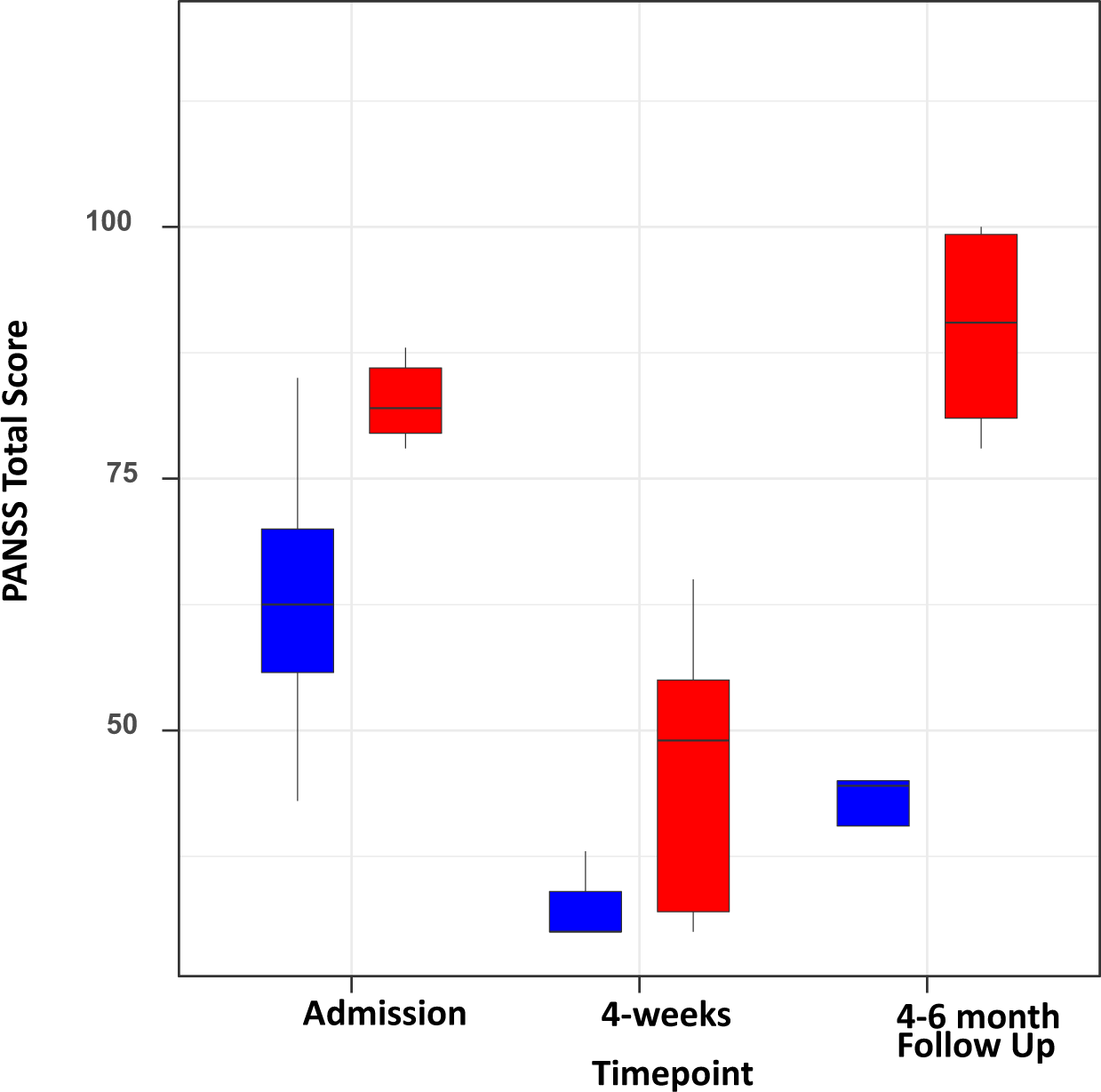
Differences in Symptoms: Relapsed vs Non-Relapsed. 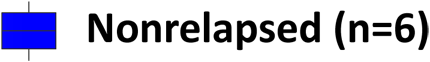 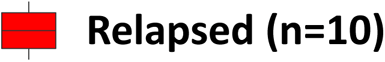 Total PANSS (Positive and Negative Syndrome Scale Scores in those who relapsed and those who did not relapse at Admission to the Hospital, at ∼ 4 weeks, and at 4-6 months. Means and +/- 2SE

## Discussion

To our knowledge, this is the first study to prospectively characterize the clinical and cognitive presentation and short-term course of CIPD and contrast it with psychosis unrelated to cannabis. Despite both groups reaching the clinical threshold for acute inpatient hospitalization for the first time as determined by independent clinicians, compared to PsyNoCan, the presentation of CIPD is characterized by significantly less severe positive, negative and general symptoms, but no significant group differences in the severity of affective symptoms. While core symptoms of psychosis improved in both groups, in contrast to the PsyNoCan group, after ∼4 weeks of inpatient treatment, the majority of the CIPD group recovered to a greater extent with minimal residual symptoms. While there were no significant group differences in cognitive function at admission, after 4 weeks of hospitalization, there was a substantial improvement in cognitive functioning in CIPD but not the PsyNoCan group. Lastly, while the relapse of psychosis in the immediate post-discharge period was temporally linked to relapse of cannabis use, the group that relapsed had greater residual psychotic symptoms at discharge.

With the liberalization of cannabis laws, commercialization of cannabis, increased access to cannabis, and greater potency of herbal cannabis and edibles, the negative consequences of cannabis exposure are expected to increase. The rates of psychoses attributable to cannabis have reportedly increased ^13, 21^, and there is a linear dose-response relationship ^22^.As the rates of CIPD are expected to increase^21^. Thus, characterizing the short- and long-term course, expression, and biology is critical to develop tailored treatments for it and to limit iatrogenic problems. For example, could chronicity be averted if the factors leading to chronicity are identified (e.g., cannabis), or are lower doses of antipsychotic drugs sufficient to treat CIPD?

While the existing literature that is based on retrospective comparisons suggests that the prominent mood symptoms but not psychosis symptoms differentiate CIPD from SCZ^23, 24^, we found no differences in affective symptoms but lower severity of positive, negative and disorganization symptoms in the CIPD group. We hypothesize that the lower severity of positive and negative symptoms might allow the affective symptoms to be more clearly observed in CIPD; alternatively, the greater severity of psychotic symptoms might obscure mood symptoms in SCZ leading to the clinical impression that CIPD presents with more affective symptoms. Furthermore, our findings on individual symptoms of PANSS corroborate with previously noted less severe impairments in insight in CIPD^23^ (Figures S5-S7). However, in contrast to the existing literature, we did not observe any group differences in depression and manic symptomatology. This was also corroborated by an independent comparison of the 5-factor model of PANSS where CIPD had lower symptoms on positive, negative and disorganization domains at baseline but comparable severity of symptoms on the excitement and emotional domains (Figure S8). The rapidity of onset of psychotic symptoms may trigger hospitalization in CIPD in contrast to PsyNoCan where onset of psychotic symptoms is more insidious.

Over the course of 4 weeks of similar treatment (the same institution, independent clinicians, treatment protocols), the CIPD group had significant reductions in psychosis symptoms; total PANSS score decreased from 73.88 (27.21) to 47.5 (17.37), a ∼40% reduction. These findings are consistent with those reported by Kullahalli that CIPD patients had a near complete resolution of psychosis after a week of abstinence from cannabis^24^. In contrast, after 4 weeks, total PANSS scores in the PsyNoCan group decreased to 74.95 (23.85), equivalent to the CIPD group at admission.

The effects of cannabis on cognitive function in individuals with psychosis are unclear. Studies comparing SCZ patients with and without cannabis use have yielded mixed findings^,25^, with some studies suggesting that cannabis using SCZ patients may have better cognitive functioning^25, 26^, even during the first episode of psychosis^26^. In the only study reporting cognitive function in patients specifically with CIPD, the CIPD group had significantly better performance on tests of intelligence and attention, perceptual tracking and sequencing compared to cannabis using SCZ patients. Furthermore, whereas cannabis using SCZ group performed worse than healthy controls across most cognitive domains, the CIPD group had no significant deficits compared to healthy controls except for non-perseverative errors in a test of executive functioning^27^. In contrast to the results of this small (n=20), cross-sectional study we find that cognitive function in CIPD is comparable to, but numerically worse than, the PsyNoCan group at the time of admission to the hospital. That cognitive test performance in the CIPD group was not significantly different from the PsyNoCan group at the time of admission may be explained by a high quantity (median 730 joints) and duration (median 8 years) of cannabis use by the CIPD group (Table 1) in our sample compared to most earlier studies. Chronic heavy cannabis use may have added to the cognitive burden^28^ imposed by the emergence of psychosis in our sample. Consistent with the explanation, a study that stratified cannabis using first episode patients by the quantity and frequency of cannabis use noted that heavy users have worse performance on cognitive tasks compared to non-users, whereas medium users didn’t show a significant difference^29^.

Cognitive function in the CIPD group improved substantially over the ∼4 weeks of hospitalization, in contrast to the PsyNoCan group. The greater improvement in the CIPD group over the 4 weeks of hospitalization may be attributed to some combination of abstinence from cannabis as well as a significant improvement in psychosis. While not part of primary aims, in order to benchmark the cognitive outcomes, and explore the independent and interactive effects of psychosis (+/-) and cannabis (+/-) on cognitive outcomes in a 2 × 2 design we also studied community-based healthy controls (n=30) without psychiatric or substance use problems from the, and individuals hospitalized for ICD-10 Cannabis Use Disorder (n=16) but without psychosis. Relative to healthy controls, both psychosis groups had significantly worse performance on cognitive outcomes at admission (Figures S9 & S10) and after 4 weeks (Table S7). Relative to the CUD group, at admission both the CIPD and PsyNoCan groups had significantly worse performance on HVLT outcomes, but only the CIPD group had lower Cogstate Schizophrenia Battery composite score (Table S7). However, after 4 weeks the CIPD group had worse HVLT scores but comparable Cogstate Schizophrenia Battery to the CUD group. Relative to the CUD group, after 4 weeks he PsyNoCan group continued to perform significantly worse on the HVLT and Groton Maze recall task. Collectively these findings suggest that CIPD may have a unique cognitive profile and course.

Of the 16 CIPD cases for whom longitudinal data was available, 10 were re-hospitalized for a relapse of psychosis related to the resumption of cannabis use, in the short follow-up period. In contrast, in the remaining 6 who did not relapse only 1 of the tested positive for cannabis. This would suggest a role for cannabis in relapse. Furthermore, in addition to cannabis use, the CIPD cases who relapsed were more symptomatic both at admission and 4 weeks compared to the those who didn’t relapse (Figure 4). Related to this, Kulahalli reported that CIPD patients presenting with nonaffective psychosis were more likely to develop a recurrent psychotic disorder than those presenting with affective psychosis^30^. Our findings that CIPD patients relapsed taken together with the literature that upto 50% of CIPD cases are “rediagnosed” schizophrenia or a related disorder in the ensuing years^9^ suggests that an initial episode of psychosis related to cannabis may be a harbinger of a recurrent psychotic disorder, that in our current nosology is lumped under schizophrenia. Why some, but not others, cases of CIPD develop a chronic recurrent psychotic disorder is not known and warrants further study.

While the study’s findings that psychosis occuring in the context of cannabis use may have a distinct initial presentation and short-term course it remains unclear is the degree to which it is distinct from other psychoses lumped under schizophrenia. Identification of biomarkers, deep phenotyping, characterization of the longitudinal course and prognosis will help disentangle the degrees of convergence and divergence across these syndromes. While admittedly speculative, the study findings raise the intriguing possibility of a subtype of schizophrenia that is “related to” or “caused” by cannabis use.

The study had some strengths and weaknesses. Other drug use relevant to psychosis (e.g., cocaine, amphetamine, phencyclidine, LSD, etc.) was non-existent in the sample lending greater clarity in interpreting the study findings. The duration of hospitalization (>4 weeks) allowed for the careful and thorough characterization of cases and controls which would hard to do in the West. The study of CUD and healthy controls, although of a smaller number, permitted disentangling the individual and interactive effects of psychosis and cannabis. Studying the groups who received the same care in the identical setting minimized other sources of variability. Given the small number of subjects in whom post-discharge longitudinal data were obtained, the follow up data analysis should be considered preliminary, and furthermore, the observed estimates for rates of relapse of psychosis in such short duration may be biased towards overestimation. Lastly, given that this study was conducted at an inpatient hospital predominantly serving males, only males were enrolled in this study. Future studies that include females will be critical in characterizing CIPD.

## Conclusions and future directions

In contrast to psychosis unrelated to cannabis, CIPD presents with less severe psychosis symptoms, comparable affective symptoms, and greater resolution of psychosis. Likewise, cognitive deficits in CIPD resolve more completely when compared to PsyNoCan. Finally, relapse of cannabis use may predict poorer long-term outcomes with greater psychotic relapses. These findings raise several questions that need to be addressed. What are the factors that contribute to the conversion to a chronic psychotic disorder? Are any of these preventable? For example, if it is known that continued use of cannabis is a significant contributor to the conversion, treatments that target cannabis use could be implemented. What is the course of the disorder? How does it respond to existing pharmacological treatments? Are there biomarkers that characterize CIPD? To what extent does it interfere with social, educational, and occupational functioning? Is the presentation and course different in females? And lastly, what is the overall prognosis?

## Data Availability

All data produced in the present study are available upon reasonable request to the authors after publication.

## Acknowledgements

Funded by the US National Institute on Drug Abuse (1R21DA041539-01 DCD), Young Investigator Award from the Brain Behavior Foundation (SG). The authors acknowledge the contributions of Ms. Renu Gehlot in collecting some study data, Ms. Christina Luddy in coordinating the study, and the late Dr. Christoday Khess for facilitating the study.

