## Supplementary material for "Presentation and Short-term Course of New Onset Cannabis Induced Psychotic Disorder in Males": s table

**Supplementary Material (in order of citation in the manuscript)**

**Table of Contents:**

1. Figure S1: General Study Design
2. Table S1: Study Groups
3. Figure S2: Consort Chart
4. Table S2: Detailed Inclusion/Exclusion Criteria Of Primary Group
5. Table S3: Detailed Inclusion/Exclusion Criteria Of Exploratory Group
6. Table S4: Detailed Schedule of Procedures - Primary Group
7. Table S5: Detailed Schedule of Procedures - Exploratory Group
8. Table S6: Detailed Outcome Measures
9. Figure S3: Chillum – Clay Pipe
10. Figure S4: Examples of Edibles
11. Figure S5: Individual PANSS Positive Item Scores At Admission
12. Figure S6 - Individual PANSS Negative Item Scores At Admission
13. Figure S7 - Individual PANSS General Item Scores At Admission
14. Figure S8 - 5 factor PANSS Psychosis At Admission
15. Figure S9. Group Differences in Verbal Memory At Admission
16. Figure S10. Group Differences in Cognitive function at Admission
17. Table S7: Differences in Admission and 4 weeks Cognitive Test Performance in CIPD and PsyNoCan compared to Healthy Controls and Cannabis Use Disorder

| **Table S1: Definition of Study Groups** | | | |
|---|---|---|---|
| | **1. Immediate Confirmation of Urine THC-COOH**<br>**2. Cannabis Use History (TimeLine Follow Back [TLFB])**<br>**3. Collateral History from Family**<br>**4. Later Quantification of Urine THC-COOH** | | |
| | **Cannabis +** | **Cannabis -** | |
| **Psychosis +** | CIPD | PsyNoCan | **Primary Sample** |
| **Psychosis -** | Cannabis Use Disorder (CUD) | Healthy Controls (HC) | **Exploratory sample** |

| Table S2: Detailed Inclusion-Exclusion Criteria of Primary Groups (CIPD & PsyNoCan) | | |
|---|---|---|
| | ***Inclusion Criteria*** | ***Exclusion Criteria*** |
| ***CIPD*** | Psychosis | Other drug exposure (except nicotine) |
| | Requiring Hospitalization | |
| | Urine THC-COOH positive | |
| | Self-reported cannabis use | |
| | Collateral reported cannabis use | |
| ***PsyNoCan*** | Psychosis | Other drug exposure (except nicotine) |
| | Requiring Hospitalization | |
| | Urine THC-COOH negative | |
| | No self-reported cannabis use | |
| | No collateral-reported cannabis use | |

| Table S3: Detailed Inclusion -Exclusion Criteria of Exploratory Sample | | |
|---|---|---|
| ***CUD*** | ICD-10 Cannabis Use Disorder | No psychosis |
| | Urine THC-COOH positive | |
| | Self-reported cannabis use | |
| | Collateral-reported cannabis use | |
| | Requiring Hospitalization | |
| ***Healthy Controls*** | No ICD10 psychiatric disorder | |
| ***Common Criteria*** | | Other drug exposure (except nicotine) |
| | | Significant neurological disorders |
| | | Significant medical disorders |

| Table S4: Schedule of Procedures - Primary Group (Hospitalized Cases of CIPD and PsyNoCan) | | | | | |
|---|---|---|---|---|---|
| | | Screen: Day 1 - 3 | Within 1 week | Discharge | 4- 6 months |
| Informed Consent | Informed Consent Process | x | | | |
| Chart review | | x | | x | |
| Demographic data | | x | | | |
| History | Initial Psychiatric Interview: JS and SS | x | | | |
| | Psychiatric Evaluation | | x | | x |
| | Cannabis Exposure History (SALCU) | | x | | x |
| | 30day TLFB (Cannabis & other Drugs) | | x | | |
| | Family history (Psychosis and Drugs) | x | | | |
| Standardized Assessments | Diagnosis (DIGS) | | x | | |
| | PANSS | x | | x | x |
| | SPQ | | x | | |
| Cognitive Testing | HVLT | | x | x | x |
| | CogState | | x | x | x |
| Psychophysiology | Resting | x | x | x | x |
| | ASSR | x | | | |
| | P300 | | x | x | x |
| Laboratory Tests | Urine Sample for Immediate Testing | x | x | x | x |
| | Urine Sample for later quantification of THC-COOH | x | x | x | x |

SALCU = Scale for the Lifetime Assessment of Cannabis Use; TLFB = Time Line Follow Back; DIGS = Diagnostic Interview for genetic studies; PANSS = Positive and Negative Syndrome Scale; SPQ = Schizotypal Personality Questionnaire

| Table S5: Schedule of Procedures - Exploratory Sample<br>(Healthy Controls and Hospitalized Cases of Cannabis Use Disorder) | | |
|---|---|---|
| Informed Consent | Informed Consent Process | x |
| Demographic data | | x |
| | Psychiatric Evaluation | x |
| | Cannabis Exposure History (SALCU) | x |
| | 30day TLFB (Cannabis & other Drugs) | x |
| | Family history (Psychosis and Drugs) | x |
| | PANSS | x |
| Cognitive Testing | HVLT | |
| | CogState | x |
| Psychophysiology | P300 and resting | x |
| Laboratory Tests | Urine Sample for Immediate Testing | x |
| | Urine Sample for later quantification of THC-COOH | x |
| SALCU = Scale for the Lifetime Assessment of Cannabis Use; TLFB = Time Line Follow Back; PANSS = Positive and Negative Syndrome Scale; | | |

### Table S6: Detailed Outcome Measures

Cannabis exposure was established with combination of self-report, collateral report from the family, a structured interview, immediate (real-time) urine toxicology and later quantification of THC-COOH in stored urine samples.

**Scale for the Lifetime Assessment of Cannabis Use (SALCU):** Cannabis use pattern was measured using the Scale Assessing Lifetime Cannabis Use (SALCU), a 27-item scale developed in our lab[1] . The SALCU comprehensively evaluates multiple domains of use pattern including age of onset, duration of use, most severe use pattern, recent use pattern, attempts to quit and lifetime cumulative cannabis exposure in standard joint equivalents.

A calendar-based approach, the **Time Line Follow Back**(TLFB)[2] was used to estimate last month drug use, including cannabis and alcohol.

Urine was tested in real time using immunoassay **PreScreen Plus Dip Card** kits on site https://drugtestsinbulk.com/ten-panel-prescreen-plus-dip-card-clia-waived.html. Testing for 10 drugs including amphetamines, barbiturates, benzodiazepines, cocaine, methadone, methamphetamines, opioids, phencyclidine, tricyclics, THC-COOH. The cut-off for detecting THC-COOH was 50 ng/dl.

**Quantification of THC-COOH in urine:** In addition, urine samples were collected at CIP, stored in a -80 ºC freezer, and transferred in large batches for analysis (Gas Chromatography Mass Spectroscopy) by the Toxicology lab, Centre for Addiction Medicine, NIMHANS, directed by Dr. Pratima Murthy and Priyamvada Sharma for cannabinoids and other drugs (opioids, benzodiazepines, cocaine, amphetamine, barbiturates, etc).

Diagnosis was established using the **Diagnostic Interview for Genetic Studies (DIGS)**[3], a structured interview schedule developed by the National Institute of Mental Health USA. It enables a careful assessment of major mood and psychotic disorders, as well as their spectrum conditions. The DIGS has been validated for psychiatric research in India[4].

Psychosis was assessed using the **Positive and Negative Syndrome Scale (PANSS)**[5] which contains subscales for positive symptoms, negative symptoms and general symptoms. The patient is rated from 1 to 7 on 30 different symptoms based on the interview as well as reports of family members or primary care hospital workers. The PANSS has been validated for use in India[6]. Based on meta-analytic results, an alternative five-factor solution of the PANSS was proposed with positive symptoms, negative symptoms, disorganization, excitement, and emotional distress.

Depression was assessed using the **Calgary Depression Rating Scale (CDRS)**[7] a 9-item structured interview scale that was designed specifically to assess depression independently of symptoms of psychosis in schizophrenia. The CDRS was developed from, and validated against, the Hamilton Depression Rating Scale, Beck Depression Inventory, and the Brief Psychiatric Rating Scale using factor analysis, internal consistency, and face validity. Items were constructed to measure: 1) Depression, 2) Hopelessness, 3) Self-deprecation, 4) Guilty ideas, 5) Pathological guilt, 6) Morning depression, 7) Early wakening, 8) Suicidal ideation; and 9) Observed depression. Items are graded on a 4-point Likert type scale (0, absent; 1, mild; 2, moderate; 3, severe), anchored by descriptors. It has been translated to Hindi and has been validated for use in India[8].

Mania was assessed using the **Young Mania Rating Scale (YMRS)**[9] a scale of eleven items based on the patient's subjective report of her/his clinical condition. The items assessed include Elevated Mood, Increased Motor Activity-Energy, Sexual Interest, Sleep, Irritability, Speech, Language-Thought Disorder, Content, Disruptive-Aggressive Behavior, Appearance, Insight. The YMRS has been validated for use in India[10].

The PANSS, CDRS and YMRS were administered by a trained Masters' level rater. Prior to initiating the study, interrater reliability sessions were conducted for all raters with the PI. Every interview was videotaped so that a random sampling of interviews could be later reviewed by a 3rd party to ensure accurate ratings.

The **CogState Schizophrenia Battery** was used to assess speed of processing, attention/vigilance, working memory, visual learning, verbal learning, reasoning/problem solving, and social cognition, domains that are recommended by the MATRICS initiative[11-14]. It is rapid (35 minutes), it uses culture-neutral stimuli (playing cards) and is computerized.

A modified **Hopkins Verbal Learning Test (HVLT)**[15,16] that has been translated for India[17] was used to measure verbal memory and recall. Learning and recall were measured using the Hopkins Verbal Learning Test (HVLT). The test consists of 3 consecutive trials of immediate free recall of a 12-item, semantically categorized list, followed 30 minutes later by testing of delayed free recall, cued recall and recognition recall.

All cognitive testing was conducted in a quiet room with the Masters' level rater present in the room.

**Table S7: Differences in Admission and 4 weeks Cognitive Test Performance in CIPD and PsyNoCan compared to Healthy Controls and Cannabis Use Disorder**

| | | | | | | | | Healthy Controls vs. | | | | Cannabis Use Disorder vs. | | | |
|---|---|---|---|---|---|---|---|---|---|---|---|---|---|---|---|
| | Test | CIPD (z score) | | PsyNoCan (z score) | | HC (z score) | CUD (z score) | CIPD (p value) | | PsyNoCan (p value) | | CIPD (p value) | | PsychNoCan (p value) | |
| | | Admission | 4 weeks | Admission | 4 weeks | Baseline | Baseline | Admission | 4 weeks | Admission | 4 weeks | Admission | 4 weeks | Admission | 4 weeks |
| HVLT - recall | Immediate | -0.51 | -0.16 | -0.18 | -0.36 | 1.08 | 0.79 | <0.0001 | <0.0001 | <0.0001 | <0.0001 | <0.0001 | <0.0001 | <0.0001 | <0.0001 |
| | Delayed | -0.49 | -0.07 | -0.28 | -0.12 | 0.99 | 0.43 | <0.0001 | <0.0001 | <0.0001 | <0.0001 | <0.0001 | 0.06 | 0.006 | 0.043 |
| | Cued | -0.72 | -0.23 | 0.004 | -0.05 | 1.16 | 0.71 | <0.0001 | <0.0001 | <0.0001 | <0.0001 | <0.0001 | 0.004 | 0.005 | 0.004 |
| | Recognition | -0.42 | -0.14 | -0.12 | -0.5 | 1.08 | 0.55 | <0.0001 | <0.0001 | <0.0001 | <0.0001 | <0.0001 | 0.008 | 0.001 | <0.0001 |
| Cogstate Schizophrenia Battery (CSB) | Composite score | **-0.26** | **0.04** | **-0.04** | **0.06** | **0.67** | **0.18** | <0.0001 | <0.0001 | <0.0001 | <0.0001 | 0.098 | 0.56 | 0.38 | 0.35 |
| | Identification | -0.16 | 0.09 | -0.17 | -0.06 | 0.77 | 0.06 | 0.006 | 0.04 | 0.007 | 0.01 | 0.52 | 0.91 | 0.52 | 0.73 |
| | Detection | -0.32 | -0.11 | 0.03 | 0.14 | 0.53 | 0.27 | 0.005 | 0.04 | 0.1 | 0.2 | 0.08 | 0.26 | 0.48 | 0.7 |
| | ONB | -0.27 | 0.22 | -0.05 | 0.06 | 0.32 | -0.1 | 0.02 | 0.69 | 0.17 | 0.3 | 0.57 | 0.31 | 0.89 | 0.6 |
| | OCL | -0.29 | 0.03 | -0.16 | 0.04 | 0.82 | 0.33 | <0.0001 | 0.002 | 0.0002 | 0.005 | 0.11 | 0.43 | 0.2 | 0.47 |
| | GML | -0.31 | 0.12 | 0.06 | -0.31 | 0.67 | -0.08 | <0.0001 | <0.0001 | 0.0006 | <0.0001 | 0.61 | 0.65 | 0.77 | 0.62 |

| | | | | | | | | | | | | | | | |
|---|---|---|---|---|---|---|---|---|---|---|---|---|---|---|---|
| | **GMR** | -0.16 | 0.2 | -0.2 | -0.39 | 0.57 | 0.43 | | 0.0001 | 0.01 | 0.0008 | 0.0003 | 0.02 | 0.27 | 0.02 | 0.007 |
| | **2BT** | -0.45 | -0.08 | -0.05 | -0.07 | 1.05 | 0.09 | | <0.0001 | <0.0001 | <0.0001 | 0.0001 | 0.19 | 0.67 | 0.73 | 0.71 |
| | **CPAL** | -0.26 | -0.31 | 0.1 | -0.06 | 0.48 | 0.23 | | 0.002 | 0.002 | 0.11 | 0.03 | 0.24 | 0.21 | 0.75 | 0.5 |
| | **SC** | -0.31 | -0.11 | 0.04 | -0.27 | 0.84 | 0.17 | | 0.0002 | 0.001 | 0.005 | 0.0003 | 0.18 | 0.43 | 0.7 | 0.22 |

I-BT: One back Test; OCL: One Card Learning; GML: Groton Maze Learning; GMR: Groton maze recall; 2-BT: Two-back test; CPAL: Continuous paired associate learning; SC: Social Cognition.
Cogstate Battery: <after correction for multiple comparisons, individual cogstate p values < 0.005 (0.05/9) were considered significant. Values transformed and centered to mean in the entire sample such that higher scores represent better performance on each task. Raw scores were used in the statistical analyses.
