## Supplementary material for "Presentation and Short-term Course of New Onset Cannabis Induced Psychotic Disorder in Males": s fig

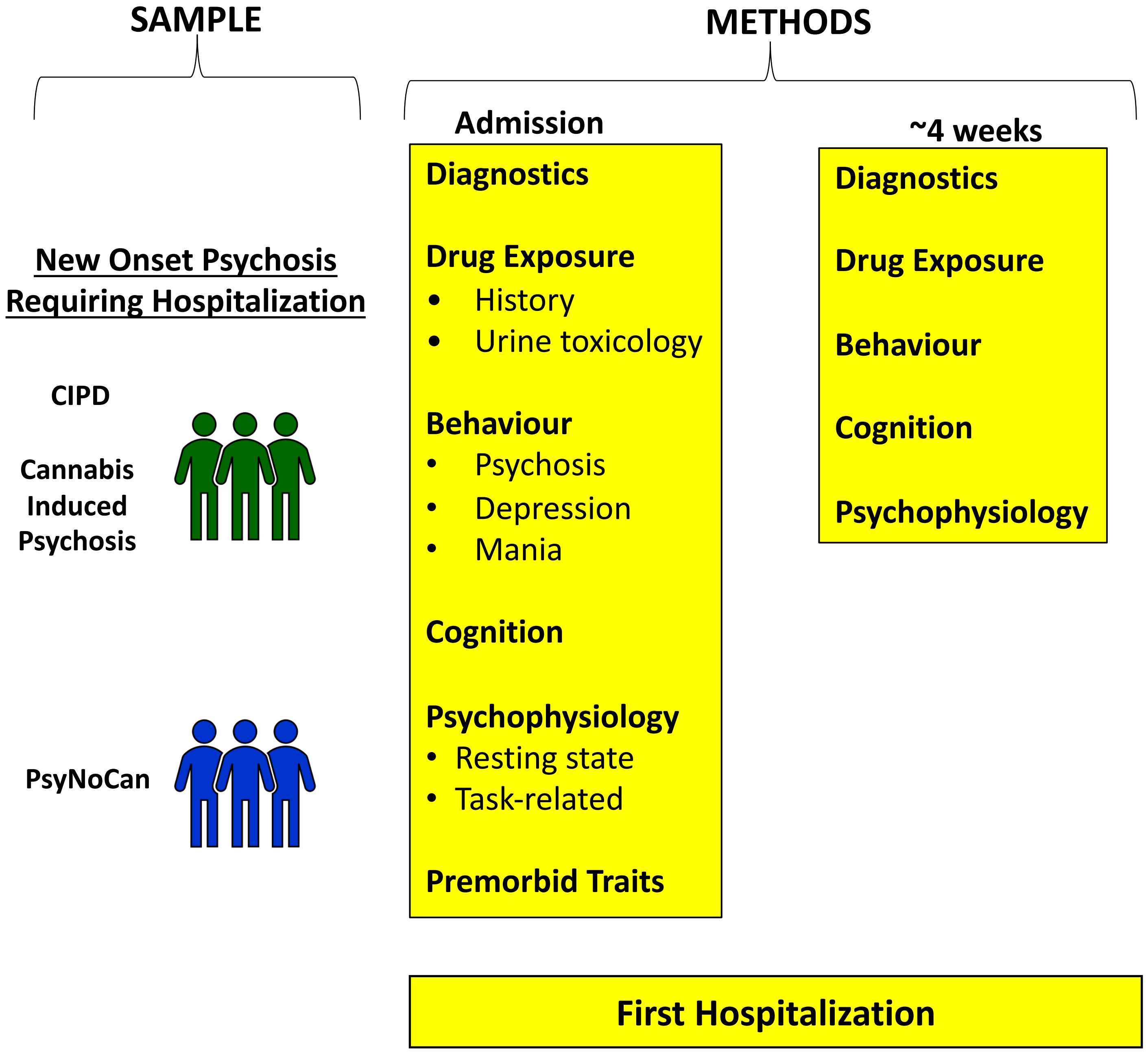


Fig S1: General Study Design

Fig S2: CONSORT chart

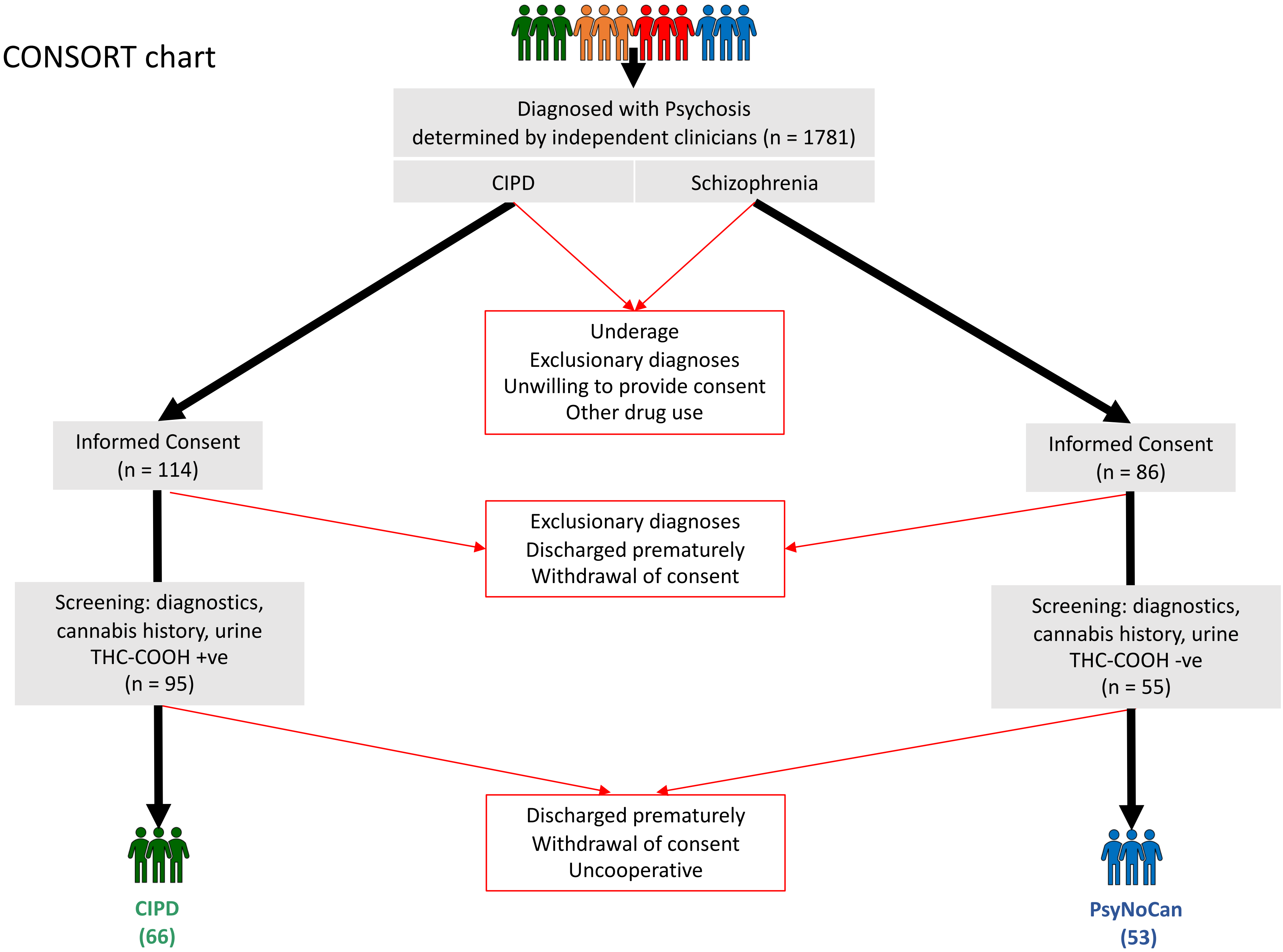

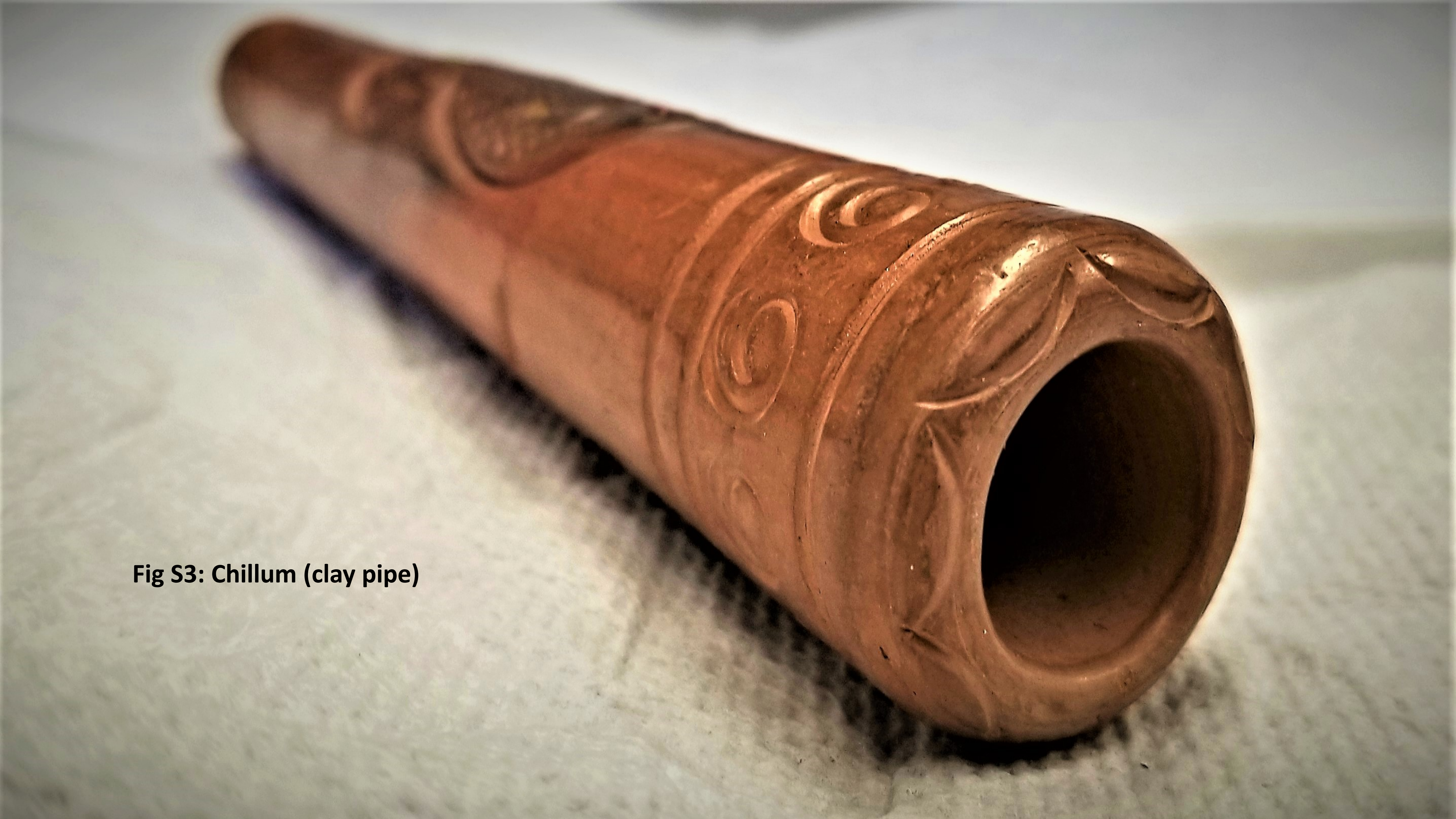

Fig S3: Chillum (clay pipe)

### Fig S4: Other modes of consumption

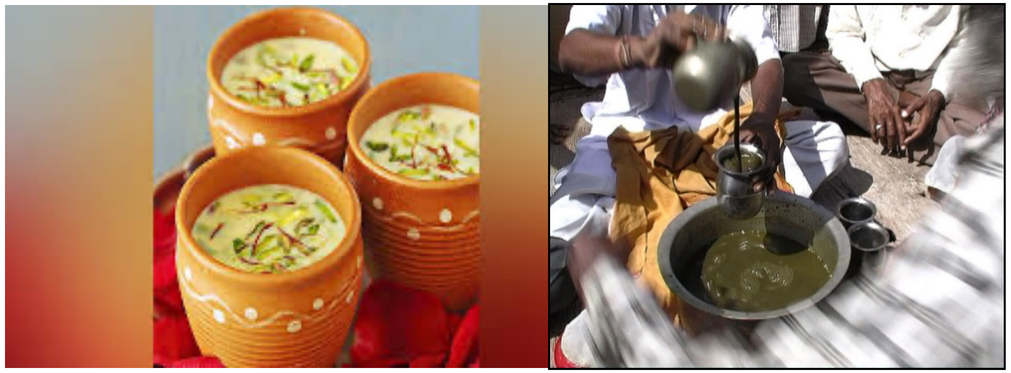

Cannabis milkshake
(“Thandai”)

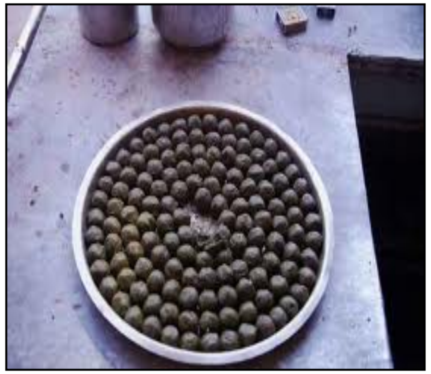

Balls of cannabis
(“Go-leez”)

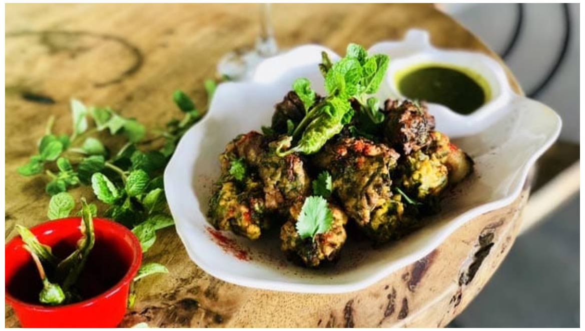

Fritters of cannabis
(“Pakoras”)

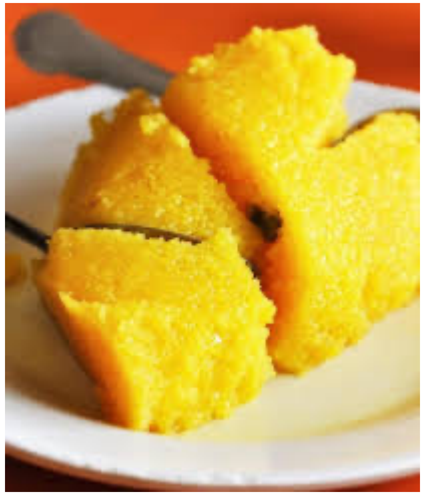

Confections of
cannabis

### Figure S5 - Individual PANSS Positive Item Scores At Admission

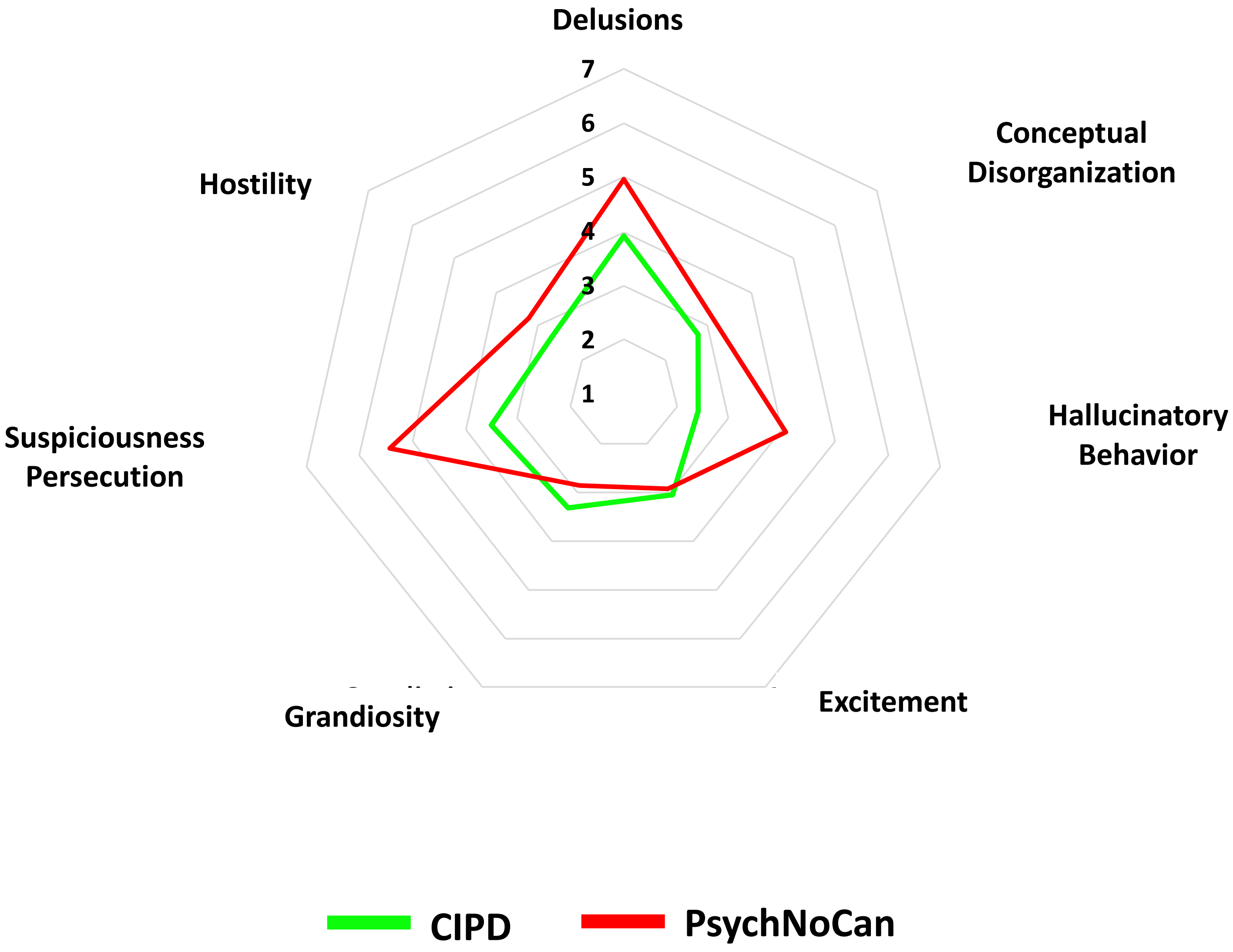

### Figure S6 - Individual PANSS Negative Item Scores At Admission

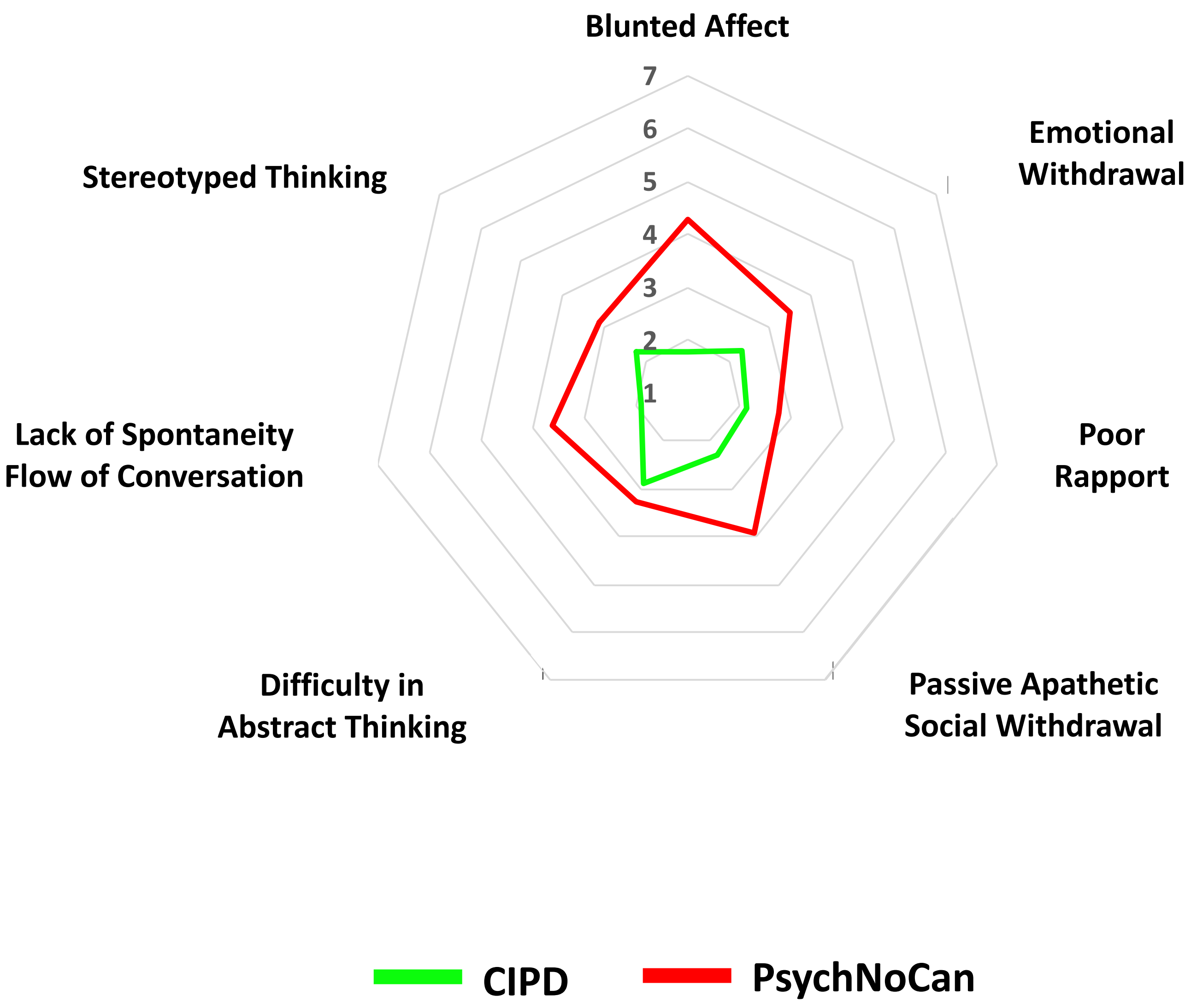

### Figure S7 - Individual PANSS General Item Scores At Admission

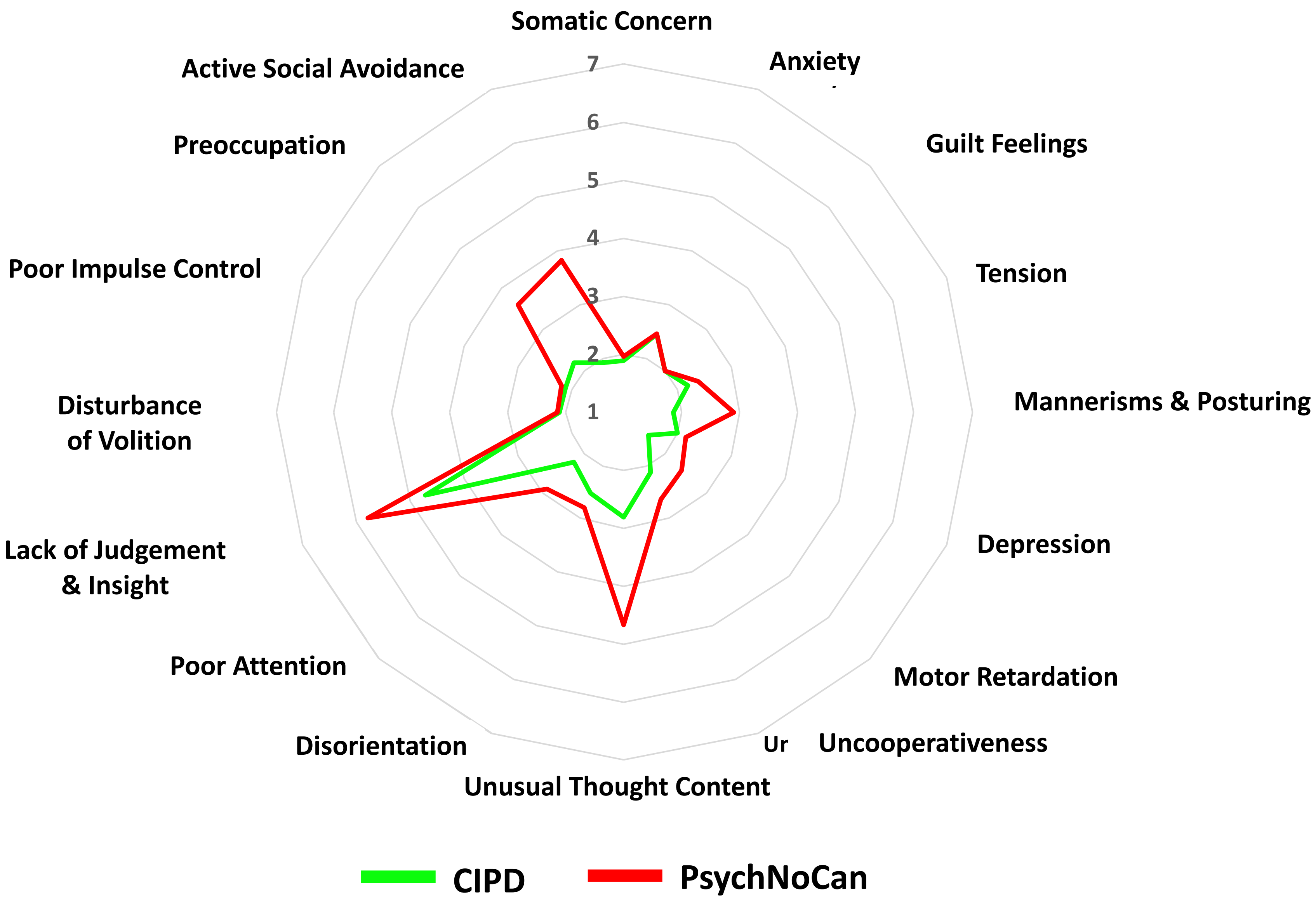

### Figure S8 - 5 factor PANSS Psychosis At Admission

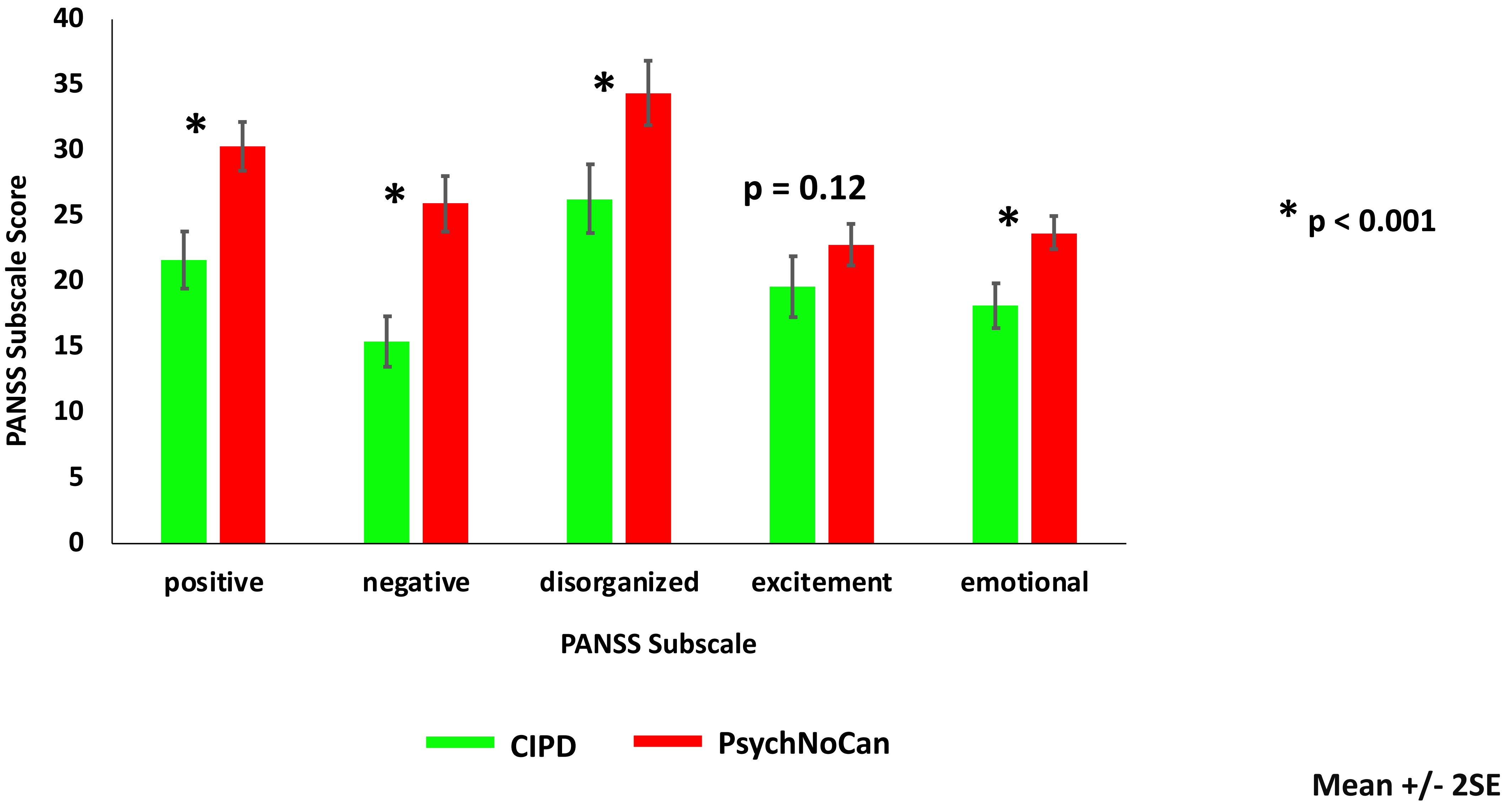

### Figure S9. Group Differences in Verbal Memory At Admission

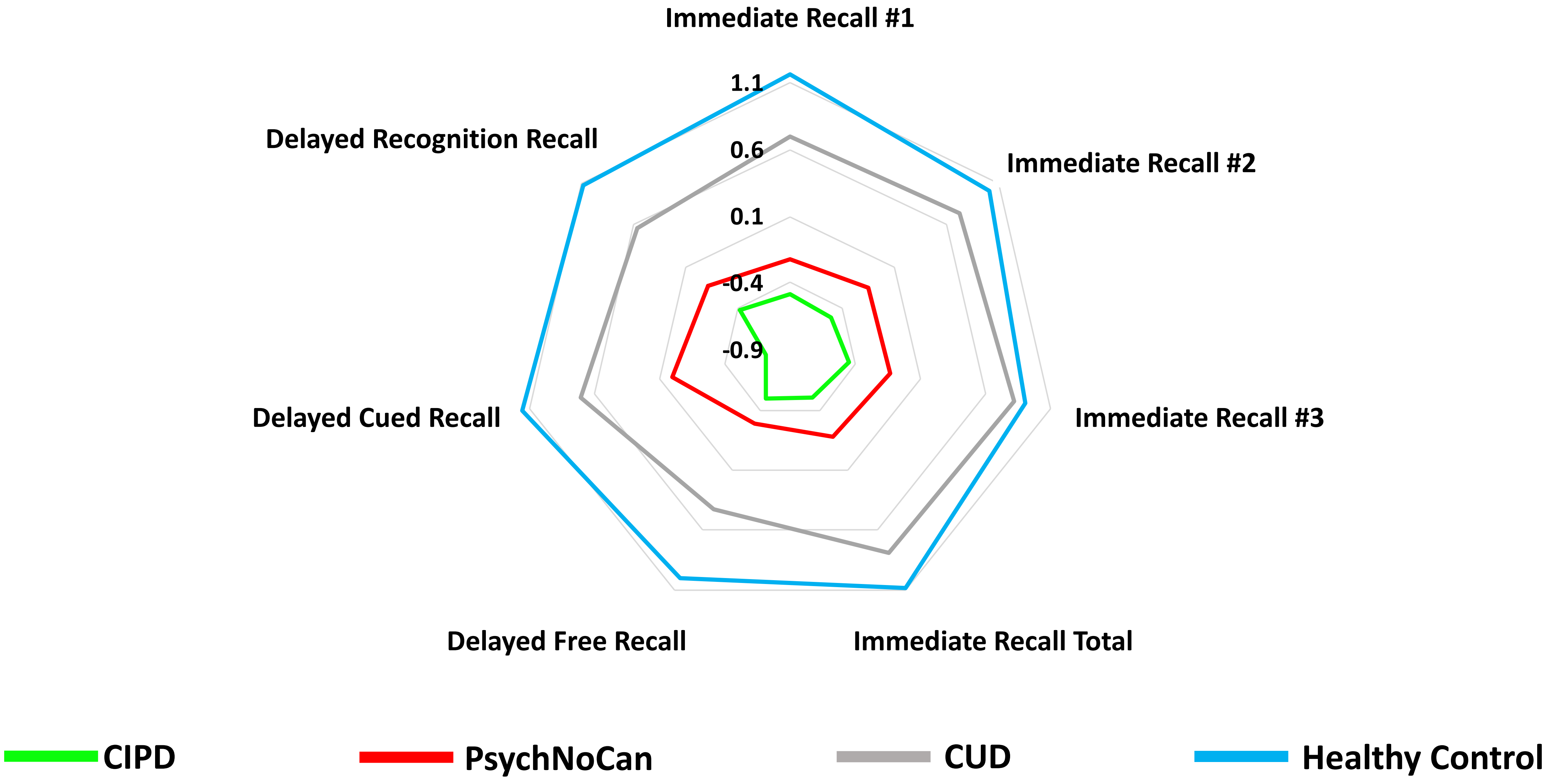

### Figure S10. Group Differences in Cognitive function at Admission

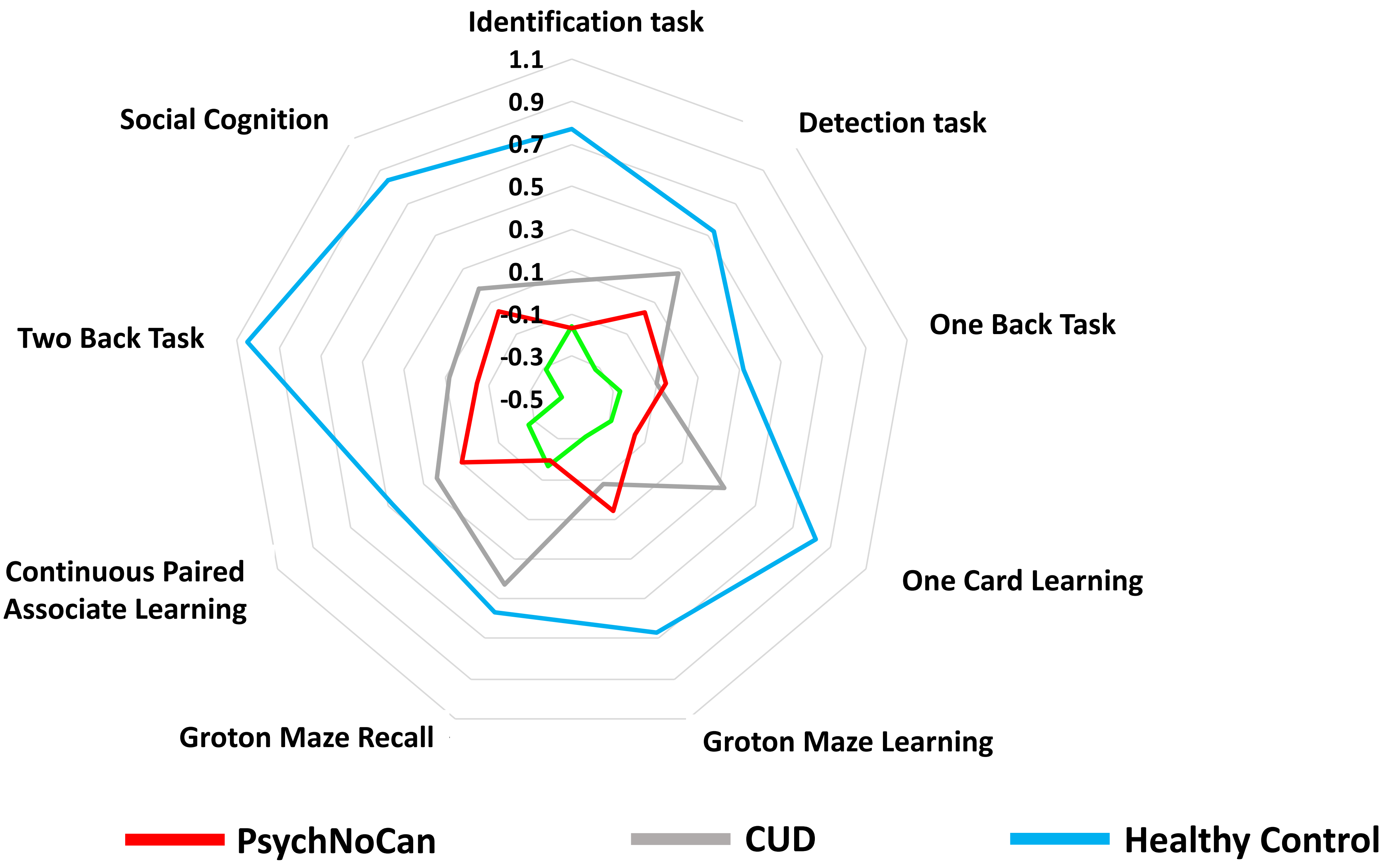
